# VAE (Variational Autoencoder) Based Gastrotype Identification and Predictive Diagnosis of *Helicobacter pylori* Infection

**DOI:** 10.64898/2026.04.11.26350690

**Authors:** Zhanshan (Sam) Ma, Yuting Qiao

## Abstract

**Background:** The enterotype concept proposed that gut microbiomes cluster into discrete types, but subsequent critiques demonstrated that such clustering depends on methodological choices, that the number of clusters is not fixed, and that faecal samples cannot capture spatial heterogeneity along the gastrointestinal tract. The stomach remains particularly understudied, and no systematic classification exists for gastric microbial community types.

**Methods:** We assembled a multi-cohort dataset of 566 gastric mucosal samples spanning healthy controls to gastric cancer, with both Helicobacter pylori (HP)-negative and HP-positive individuals. Critically, we applied the key methodological lessons of the enterotype debate: we used a variational autoencoder (VAE) for dimensionality reduction to learn a continuous latent representation without forcing discrete structure, determined the optimal number of clusters using the Silhouette index (an absolute validation measure) across K=2 to K=10 rather than arbitrarily selecting a cluster number, and performed transparent evaluation of multiple clustering solutions. This VAE-plus-silhouette workflow directly addresses the critiques leveled against the original enterotype analysis.

**Results:** Four gastotypes were identified, with K=4 achieving the highest mean silhouette score, indicating good cluster cohesion and separation. Two gastotypes (Variovorax-type and Trabulsiella-type) were significantly enriched in HP-positive samples, while two gastotypes (Bacteroides-type and Streptococcus-type) were significantly enriched in HP-negative samples. Random Forest and Gradient Boosting achieved excellent baseline performance for predicting HP infection (AUC = 0.990 and 0.993).

**Conclusions:** The VAE-plus-silhouette workflow provides a robust, data-driven approach for identifying gastotypes without forcing discrete structure or arbitrarily fixing cluster numbers. Using this framework, we identified four gastotypes with significantly different HP infection rates. Variovorax-type and Trabulsiella-type showed strong HP-positive enrichment, while Bacteroides-type and Streptococcus-type showed strong HP-negative enrichment. These findings demonstrate that methodological advances from the enterotype controversy can be successfully transferred to the stomach, offering a reproducible taxonomy for stratifying HP infection status with potential clinical utility.

## 1. Introduction

The human gastrointestinal tract harbors one of the most complex and densely populated microbial ecosystems known, with profound implications for host health, immune development, and disease susceptibility (McCallum & Tropini, 2024). For nearly two decades, researchers have sought to simplify this complexity by identifying recurrent compositional patterns that could stratify individuals into discrete microbial community types. The most influential attempt was the proposal of “enterotypes” — three robust clusters of faecal microbial communities driven by *Bacteroides*, *Prevotella*, or *Ruminococcus* (Arumugam et al., 2011). This concept was immediately appealing because it offered a categorical framework analogous to blood groups, raising the possibility of microbiome-based diagnostics and personalized therapies.

However, the enterotype concept was rapidly met with substantive critique. Jeffery, Claesson, O’Toole, and Shanahan (2012) argued that the term smuggled in unwarranted assumptions of discreteness, stability, and exclusivity, and they proposed that gut microbial variation follows continuous gradients rather than discrete clusters. Koren and colleagues systematically demonstrated that enterotype detection depends heavily on methodological choices, including the distance metric, clustering algorithm, validation statistic, and even the specific region of the 16S rRNA gene sequenced (Koren et al., 2013). When absolute validation measures such as Prediction Strength or Silhouette index were applied, support for discrete gut enterotypes was weak to moderate at best, with silhouette scores failing to reach the threshold for strong clustering (Koren et al., 2013). Knights and colleagues delivered the most direct empirical refutation, showing that a single healthy individual sampled daily for one year could traverse multiple putative enterotypes, and that a classifier using raw taxon abundances substantially outperformed one using enterotype cluster labels for predicting obesity and Crohn’s disease (Knights et al., 2014).

A major reconciliation followed. Costea and colleagues, in a paper co-authored by 32 researchers including both original proponents and critics, concluded that the gut microbial composition landscape has local optima or preferred community compositions but not discrete boundaries (Costea et al., 2018). They demonstrated that *Bacteroides* abundance follows a continuous log-normal distribution, while *Prevotella* shows bimodal distribution in some populations, and that 16% of individuals switched enterotypes over six months. The field converged on a consensus: enterotypes are useful as a heuristic for coarse population stratification, but they are not discrete biological kinds, and they cannot replace direct analysis of microbial species and functions. A comparable principle has also been observed in other host-associated microbiomes. For example, in the vaginal microbiome, community states may reflect both phenotype and community complexity rather than a single disease-aligned canonical configuration, while still remaining useful for classification and stratification (Ma & Li 2017; Ma, 2023).

Despite this hard-won methodological clarity, one critical limitation remained unresolved. Enterotypes were defined exclusively using faecal samples, which represent only the luminal content of the distal colon. Yet the gastrointestinal tract is a spatially structured ecosystem with marked gradients of pH, oxygen, transit time, mucus thickness, and immune factors along both its longitudinal and transverse axes (McCallum & Tropini, 2024; Siezen & Kleerebezem, 2011). The stomach, in particular, presents a uniquely challenging environment: low pH (1.5–3.5), a thick mucus layer, and historically low reported bacterial abundance.

A complete understanding of the gastric microbiome, however, is impossible without accounting for *Helicobacter pylori* (*H. pylori*). This bacterium colonizes the stomach of approximately half of the world’s population and is the primary risk factor for chronic gastritis, peptic ulcers, and gastric adenocarcinoma (Malfertheiner et al., 2017). *H. pylori* does not merely add one taxon to an otherwise stable community; it fundamentally restructures the entire gastric microbial ecosystem. HP-positive individuals consistently show reduced microbial diversity, altered pH profiles, and compositional patterns dominated by *Helicobacter* itself, whereas HP-negative stomachs are often enriched with oral-associated taxa such as *Streptococcus*, *Prevotella*, and *Veillonella* (Klymiuk et al., 2017; Xiao & Ma, 2022; Liu et al., 2022). Thus, any study of the gastric microbiome that ignores *H. pylori* status risks conflating two fundamentally different microbial ecosystems. Despite this, many published gastric datasets either exclude HP-positive samples entirely or lack sufficient HP-negative controls, leaving the field with an incomplete picture.

The present study addresses this gap by assembling a large, multi-cohort dataset that includes both HP-negative and HP-positive gastric mucosal samples across the full histological spectrum from healthy controls to gastric cancer (Coker et al., 2018; Ling et al., 2019; Park et al., 2019; Wang et al., 2020). This comprehensive design allows us to characterize the gastric microbiome in a manner that explicitly accounts for *H. pylori* infection status, rather than treating it as an unmeasured confounder.

In this study, we apply the key lessons of the enterotype literature — data-driven determination of cluster number, absolute cluster validation using silhouette scores, and transparent evaluation of multiple clustering solutions — to the gastric mucosal microbiome. Using a variational autoencoder (VAE) for dimensionality reduction followed by K-means clustering, we identify recurrent compositional patterns in this large, multi-cohort dataset. This gastric application was motivated in part by our recent VAE-based methodological work on microbiome community typing, which showed that latent-space learning can improve the robustness and interpretability of community-state inference while also providing a principled bridge between unsupervised stratification and supervised prediction (Qiao & Ma, 2026). We term these patterns **gastotypes** (from Greek *gaster*, stomach), distinguishing them from faecal-derived enterotypes while adhering to the rigorous clustering framework that the enterotype debate established. We then evaluate the association of each gastotype with *H. pylori* infection status and assess whether gastotype information improves predictive diagnosis of infection. By focusing on the stomach with a dataset that spans both HP-negative and HP-positive individuals across all histological stages, we demonstrate that the methodological advances from the enterotype controversy can be successfully transferred to a new anatomical site, generating biologically interpretable and clinically relevant community types.

## 2. Materials and Methods

### 2.1 Datasets of Gastric Microbiomes

Publicly available gastric mucosal microbiome datasets were collected from published 16S rRNA sequencing studies reporting both histological stage and *Helicobacter pylori* (HP) infection status. In total, 11 stage-specific cohorts were included, covering healthy controls, chronic gastritis, superficial gastritis, atrophic gastritis, intestinal metaplasia, and gastric cancer. These cohorts were derived from four published studies and included the datasets SRP200169, ERP108962, SRP100168, SRP109017, and SRP172818, corresponding to studies by Coker et al., Ling et al., Park et al., and Wang et al. (Coker et al., 2018; Ling et al., 2019; Park et al., 2019; Wang et al., 2020). All samples were gastric mucosal specimens. The final dataset comprised HP-negative and HP-positive samples from China and South Korea, and details are provided in Table S1.

Raw reads were processed using Trimmomatic (v0.39), followed by taxonomic classification with Kraken2 (v2.1.2) and Bracken (v2.6). Data conversion and abundance-table processing were performed in R (v3.6.3) with KrakenTools. Downstream analyses were conducted on genus-level OTU abundance tables obtained by collapsing the classified features to the genus level. To reduce phenotype misclassification, samples with discordant HP status between source metadata and sequencing-based taxonomic profiles were removed following the strategy of Xiao and Ma (2022). Specifically, samples annotated as HP-negative were excluded if the relative abundance of *H. pylori* exceeded 2%, whereas samples annotated as HP-positive were removed if the relative abundance of *H. pylori* was below 2%. The 2% threshold was chosen based on the recommendation of Xiao and Ma (2022), who demonstrated that this cutoff optimally balances sensitivity and specificity for HP status classification from 16S rRNA data. To achieve sufficient statistical power and coverage across the full histological spectrum, we pooled publicly available gastric mucosal 16S rRNA datasets. This multi-cohort design also enhances the generalizability of any identified gastotypes beyond single-study artifacts.

### 2.2 VAE architecture and optimization objective

Let 𝑥 ∈ ℝ*^D^* denote the transformed microbiome feature vector for a given sample. The encoder maps 𝑥to the parameters of a diagonal Gaussian variational posterior,

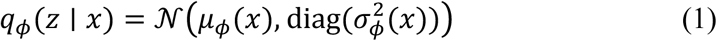

where both 𝜇_ϕ_(𝑥) and σ_ϕ_^2^(𝑥) are generated by a feed-forward neural network. Latent variables were obtained using the reparameterization trick,

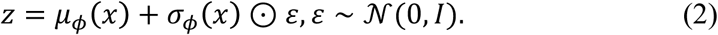

The decoder specifies 𝑝_θ_(𝑥 ∣ 𝑧)and reconstructs the input as 𝑥. Model fitting was based on a 𝛽-regularized evidence lower bound (ELBO),

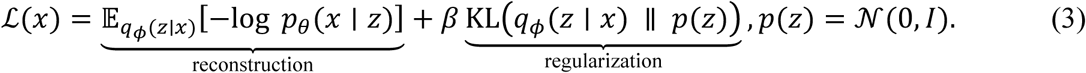

where 𝑝(𝑧) = 𝒩(0, 𝐼) . This objective function jointly optimized reconstruction fidelity and latent-space regularization, thereby generating a compact representation suitable for downstream clustering and classification. In all downstream analyses, the posterior mean of the encoder was used as the deterministic latent embedding, consistent with both the general VAE literature and our recent VAE-based microbiome community-typing framework (Kingma & Welling, 2014; Higgins et al., 2017; Qiao & Ma, 2026).

We implemented a fully connected VAE with a two-dimensional latent layer. A two-dimensional latent space was chosen to facilitate visualization and interpretation of the resulting gastotypes in a planar embedding, while retaining major variation in the data for downstream clustering (Figure 2). The encoder contained two dense hidden layers and output the latent mean and log-variance vectors, 𝜇_ϕ_(𝑥)and logσ_ϕ_^2^(𝑥). For all downstream analyses, we used the posterior mean as the sample-level embedding, i.e., 𝑧 ≡ 𝜇_ϕ_(𝑥), rather than drawing stochastic samples from 𝑞_ϕ_(𝑧 ∣ 𝑥). This deterministic representation was chosen to improve reproducibility across repeated resampling and cross-cohort evaluation and to facilitate stable nearest-centroid assignment for held-out samples (Qiao & Ma, 2026).

### 2.3 VAE-based gastrotype identification

For gastrotype analysis, taxa were retained if they had a total count of at least 5 across the dataset and occurred in at least 10% of samples. For preprocessing, genus-level count data were first converted to relative abundances, followed by Hellinger transformation through square-root scaling. The transformed matrix was then standardized using StandardScaler to obtain z-scored features. Any missing or non-finite values, including positive and negative infinities, were replaced with zero. The final processed matrix was subsequently used for variational autoencoder (VAE) training and downstream K-means clustering. Conceptually, this framework is analogous to the enterotype concept originally proposed for the gut microbiome, in which complex microbial variation can be summarized into recurrent community configurations with distinct ecological signatures (Arumugam et al., 2011). However, because the present study focused on gastric mucosal communities rather than intestinal microbiota, the term gastrotype is more appropriate than enterotype.

A variational autoencoder (VAE) was used to learn a low-dimensional representation of the gastric microbiome. The model employed a two-dimensional latent space and two fully connected hidden layers of 256 and 128 units. The encoder parameterized the mean and log-variance of a diagonal Gaussian posterior, and the decoder reconstructed the input abundance profile. Model training minimized a 𝛽-regularized evidence lower bound, with 𝛽 = 0.2. Optimization was performed with Adam (learning rate 1×10^-3^) for up to 200 epochs, with a batch size of 64, a validation split of 15%, and early stopping with a patience of 20 epochs. The *β*-value of 0.2 was selected to prioritize reconstruction fidelity over strict latent space regularization, following the general *β*-VAE framework of Higgins et al. (2017), because microbiome data contain genuine biological variation that should not be forced into a fully disentangled or perfectly Gaussian latent structure (Higgins et al., 2017).

Optimization was performed with Adam (learning rate 1×10^-3^) for up to 200 epochs, with a batch size of 64, a validation split of 15%, and early stopping with a patience of 20 epochs. The posterior mean of the encoder was used as the deterministic latent representation for downstream analyses.

K-means clustering was applied to the latent representations to identify microbial community types. Candidate cluster numbers from 𝐾 = 2 to 𝐾 = 10were evaluated, with 30 repeated k-means runs for each 𝐾. Solutions were retained only if each cluster contained at least 10 samples or at least 5% of the cohort. For each valid 𝐾, clustering quality was assessed using the mean silhouette coefficient, its standard deviation across runs, the average pairwise adjusted Rand index, and a cluster-balance metric.

Associations between gastrotype and HP infection status were assessed using chi-square tests for the overall contingency table and two-sided Fisher’s exact tests for each gastrotype versus all remaining samples. Multiple testing correction was performed using the Benjamini–Hochberg procedure. For descriptive interpretation, representative genera within each gastrotype were ranked according to mean relative abundance and fold enrichment relative to the global mean profile.

Unlike the original enterotype analysis, which arbitrarily selected three clusters based on the Calinski-Harabasz index—a relative measure that assumes clusters exist—we determined the optimal number of clusters using the Silhouette index, an absolute validation measure that quantifies how similar each sample is to its own cluster compared to other clusters (Koren et al., 2013; Rousseeuw, 1987).

### 2.4 Benchmark diagnosis with conventional machine-learning models

To establish baseline diagnostic performance, genus-level abundance matrices from HP-negative and HP-positive samples were aligned and merged into a single feature matrix, with labels coded as 0 for HP-negative and 1 for HP-positive samples. Six conventional machine-learning models were evaluated: logistic regression (LR), Random Forest (RF), k-nearest neighbors(KNN), gradient boosting (GB), linear support vector machine (SVM), and a feed-forward neural network (NN). These six models were selected to represent diverse algorithmic families: linear (LR, Linear SVM), tree-based (RF, GB), distance-based (KNN), and neural network (NN) approaches, allowing comparison of their relative strengths on high-dimensional, sparse microbiome data.

Performance was assessed using repeated stratified cross-validation with 5 folds and 20 repeats. Within each split, features were standardized using StandardScaler fitted on the training data and applied to the test data. Logistic regression was implemented with the liblinear solver, k-nearest neighbors used 𝑘 = 17, and the linear SVM used an L2 penalty. The neural network consisted of two hidden layers with 64 and 32 units and a dropout rate of 0.3, followed by a sigmoid output layer. It was trained with Adam and binary cross-entropy loss for up to 300 epochs, with 15% of the training data used for validation and early stopping with a patience of 25 epochs. Accuracy and area under the receiver operating characteristic curve (AUC) were recorded for each resampling iteration.

### 2.5 Prediction of *Helicobacter pylori* infection using gastrotype information and VAE-derived representations

To further evaluate the predictive value of gastrotype information and VAE-derived latent features, four Random Forest-based models were compared under the same repeated stratified 5-fold, 20-repeat cross-validation framework: RF using the original abundance matrix alone, Gastrotype + RF using abundance plus one-hot gastrotype labels, VAE + RF using latent features alone, and VAE + Gastrotype + RF using latent features combined with one-hot gastrotype labels. Gastrotype assignments were imported from an external annotation table and encoded as four-category one-hot variables.

For the VAE-based branches, abundance features were standardized within each training split, and a separate VAE was trained using training data only. This prediction-stage VAE used an eight-dimensional latent space, hidden layers of 128 and 64 units, and 𝛽 = 0.2, and was trained with Adam (learning rate 1×10^-3^) for up to 200 epochs, with a batch size of up to 64, a validation split of 15%, and early stopping with a patience of 20 epochs. The posterior mean of the encoder was used as the latent feature vector. Model performance was summarized by accuracy and AUC across all resampling runs, and 95% confidence intervals were estimated using the 𝑡-distribution.

The same VAE framework was used in both the unsupervised and supervised branches, but the latent dimensionality was adapted to the analytical objective. This study applied our recently developed VAE-based microbiome methodology, in which latent representations are used to connect unsupervised community typing with supervised prediction within a unified analytical framework, here adapted from gut enterotyping to gastric microbiome stratification and HP prediction (Qiao & Ma, 2026). A two-dimensional latent space was used in the unsupervised branch to facilitate visualization and interpretable gastrotype clustering, whereas an eight-dimensional latent space was adopted in the prediction branch to retain more information from the original abundance profiles for classification-oriented feature extraction. The higher-dimensional latent representation in the prediction branch was used to reduce compression-related information loss relative to the two-dimensional embedding used primarily for visualization.

## 3. Results

### 3.1 VAE-derived gastrotypes reveal four gastric mucosal communities

Whereas the original enterotype framework identified three clusters using the Calinski–Harabasz index, our analysis evaluated cluster number with the silhouette index, which emphasizes the degree of within-cluster cohesion and between-cluster separation (Rousseeuw, 1987; Koren et al., 2013). Among the tested clustering solutions, K = 4 achieved the highest mean silhouette score and was therefore selected as the optimal number of gastrotypes for downstream analyses (Figure 1 and 2, Table S2). Table S2 presents the classification of each of the 566 stomach microbiome samples as determined by our VAE-based approach. Our finding indicates that the four-cluster solution provided the strongest overall separation structure in the gastric mucosal microbiome dataset, with a more favorable balance between within-cluster cohesion and between-cluster separation than the alternative *K* values tested.

**Figure 1.**
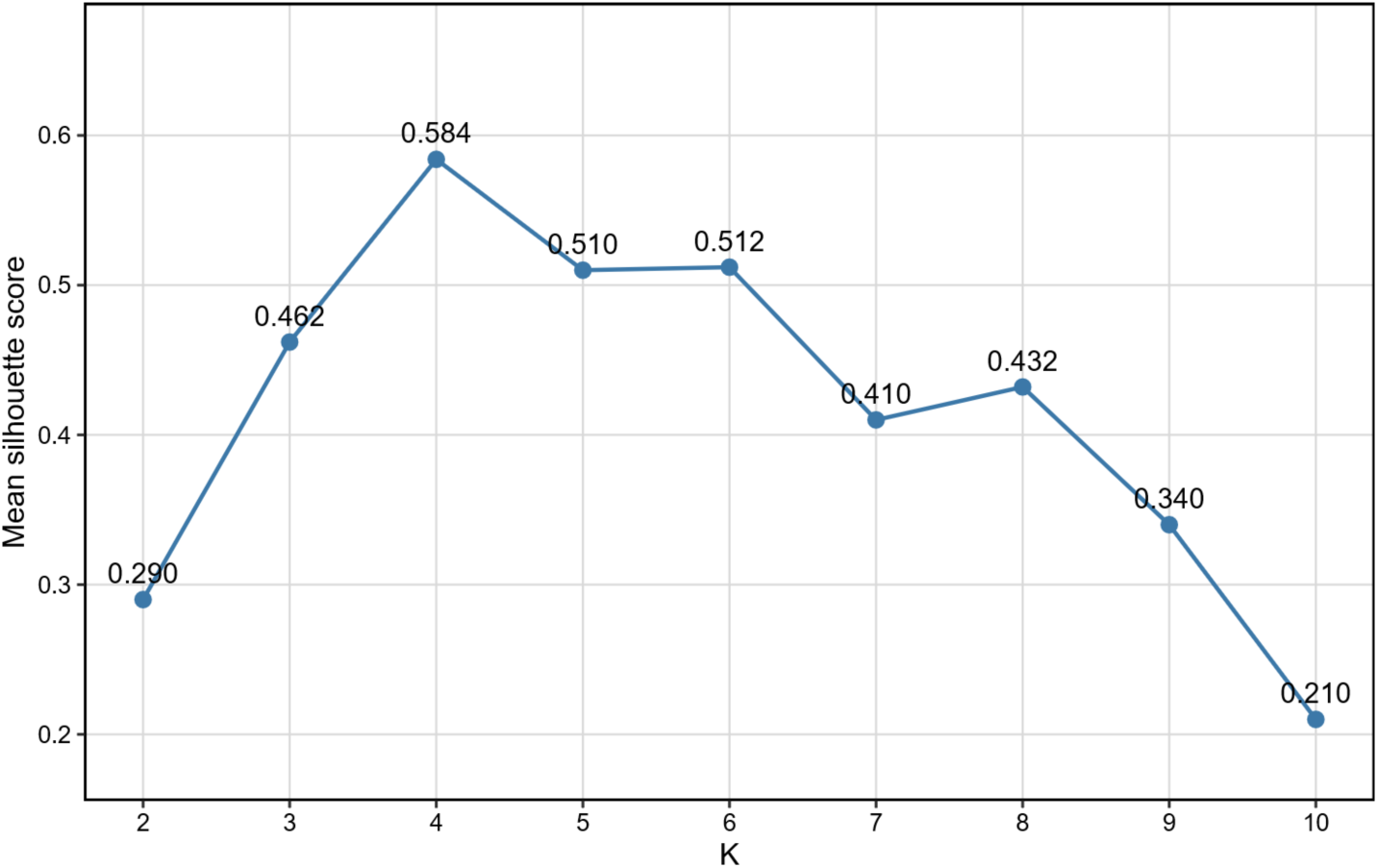
Mean silhouette scores across different K values for gastric microbial community clustering. The plot shows the mean silhouette score obtained for clustering solutions with K ranging from 2 to 10. Higher silhouette scores indicate better separation and cohesion of the inferred clusters. Among the tested values, the clustering solution with K = 4 achieved the highest mean silhouette score and was therefore selected for downstream analysis.

**Figure 2.**
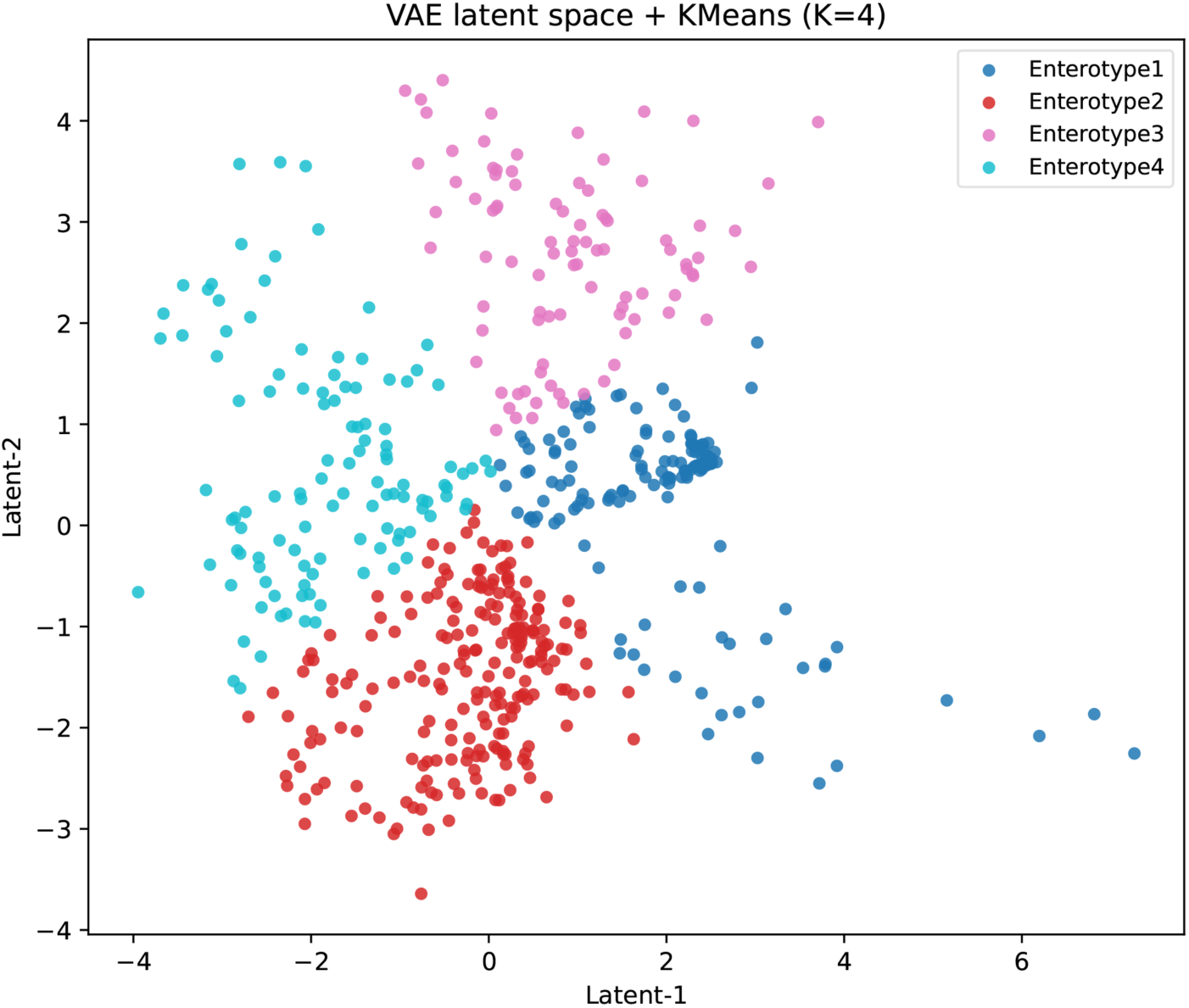
VAE latent-space representation of four gastric types identified by K-means clustering in the HP dataset. Each point represents one sample projected into the two-dimensional latent space learned by the variational autoencoder (VAE). Samples were clustered using K-means with K = 4, and colors indicate gastric type assignment. The plot shows spatial separation and relative overlap among the four gastric types in the learned latent representation. That is, the gastotypes are not absolutely discrete categories—consistent with the enterotype reconciliation (Costea et al., 2018).

**Figure 3.**
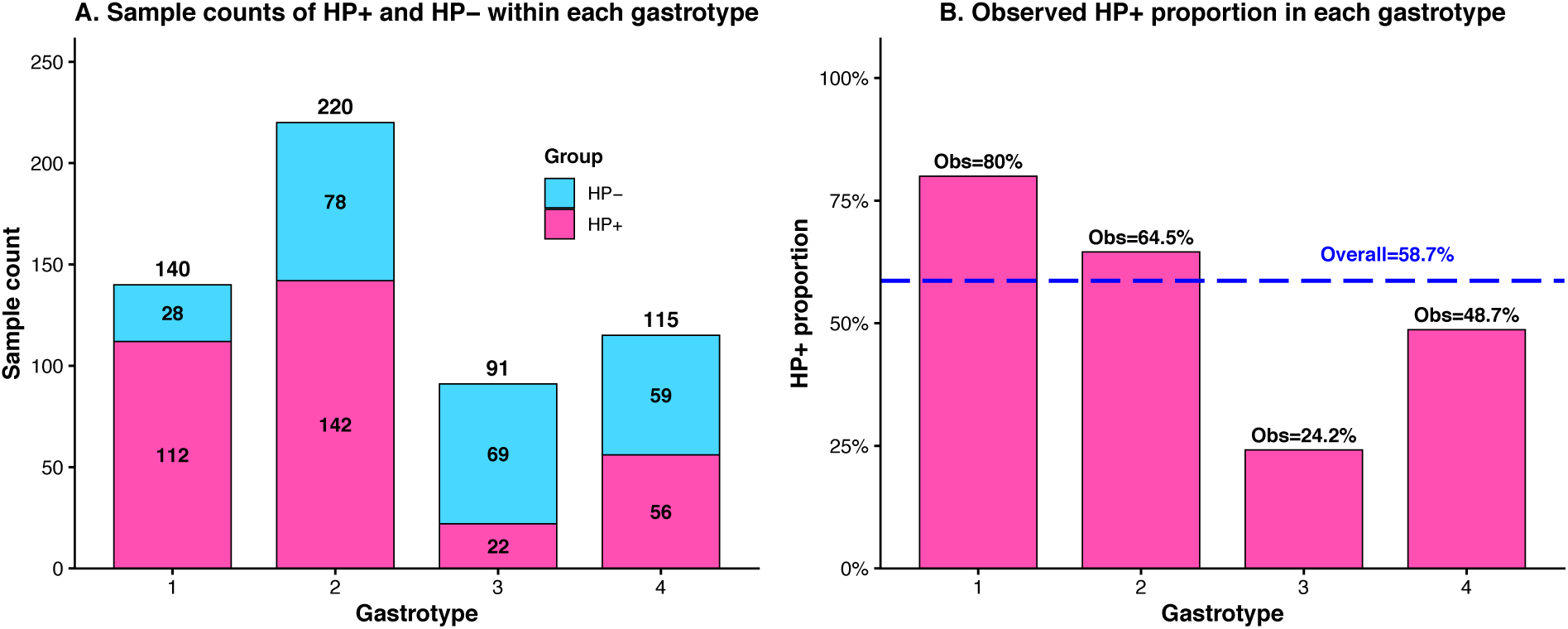
Proportion of HP-positive and HP-negative samples across four gastric type clusters. **(A)** Stacked bar plot showing the sample counts of HP− and HP+ individuals within each gastric type. **(B)** Bar plot showing the observed proportion of HP+ individuals in each gastric type. The horizontal dashed line indicates the overall HP+ proportion in the full dataset (58.7%), and the labels above the bars indicate the observed HP+ proportion for each gastric type. Together, these panels illustrate the unequal distribution of HP infection status across the four gastric types.

#### Taxonomic characterization and naming of the four gastric types

Taxonomic characterization and naming of the four gastric types were performed for descriptive and interpretive purposes using predefined heuristic criteria applied to genus-level relative-abundance profiles. For each cluster, the mean relative abundance of each genus within that cluster was compared with its overall mean relative abundance across all samples to calculate an enrichment ratio. Provisional cluster labels were then assigned by examining the three genera with the highest enrichment ratios. A genus was selected as the representative label when its mean relative abundance within the cluster was at least twofold higher than the overall mean across all samples (fold change > 2 relative to global mean) and reached at least 1% within the cluster. If no enriched genus satisfied both criteria, the label was assigned according to the genus with the highest mean relative abundance in that cluster. These labels were intended as descriptive summaries of dominant cluster features rather than formal taxonomic or ecological definitions. Compared with naming based solely on the most abundant genus, this rule-based strategy better captures cluster-specific taxonomic features by jointly considering relative enrichment and representative abundance, thereby reducing the influence of highly prevalent but weakly discriminative genera.

Gastric type 1 was designated the Variovorax-type, because *Variovorax* showed the strongest characteristic enrichment in this cluster (FC = 3.19, mean relative abundance = 0.028). In terms of overall abundance structure, this gastric type was dominated by *Helicobacter* (0.572), followed by *Pseudomonas* (0.081) and *Prevotella* (0.034), while the main enriched taxa included *Variovorax*, *Helicobacter*, and *Janthinobacterium*. These results suggest that gastric type 1 represents a *Helicobacter*-rich configuration with additional enrichment of *Variovorax*-associated signals.

Gastric type 2 was designated the Trabulsiella-type, as *Trabulsiella* was identified as a representative enriched genus in this cluster (FC = 2.39, mean relative abundance = 0.014). The most abundant genera in this gastric type were *Haemophilus* (0.136), *Helicobacter* (0.106), and *Halomonas* (0.095), whereas the principal enriched taxa included *Candidatus Portiera*, *Trabulsiella*, and *Psychrobacter*. Thus, gastric type 2 appeared to be characterized by a mixed abundance structure centered on *Haemophilus* and *Helicobacter*, with cluster-specific enrichment of *Trabulsiella*-related taxa.

Gastric type 3 was designated the Bacteroides-type, because *Bacteroides* showed the highest cluster-specific enrichment (FC = 4.91, mean relative abundance = 0.090) and was also one of the dominant genera in this cluster. In addition, this gastric type showed high mean abundances of *Faecalibacterium* (0.205) and *Prevotella* (0.151), while the top enriched taxa were *Bacteroides*, *Blautia*, and *Propionibacterium*. Collectively, these features indicate that gastric type 3 corresponds to a gut-associated commensal-like community configuration enriched in anaerobic taxa (Arumugam et al., 2011).

Gastric type 4 was designated the Streptococcus-type, as *Streptococcus* displayed both strong cluster-specific enrichment (FC = 3.78, mean relative abundance = 0.137) and the highest mean abundance within this cluster. Other dominant genera included *Lactobacillus* (0.130) and *Prevotella* (0.127), while the major enriched taxa were *Streptococcus*, *Veillonella*, and *Neisseria*. Therefore, gastric type 4 represents an oral-like mucosal community structure characterized by enrichment of *Streptococcus*-associated taxa.

The ecological characteristics of the four gastrotypes suggest biologically meaningful differentiation rather than purely mathematical partitioning. Gastrotype 1 was dominated by *Helicobacter* and therefore most likely represents a canonical HP-associated gastric state. Gastrotype 2 retained a mixed structure enriched in *Haemophilus*, *Helicobacter*, and *Halomonas*, suggesting that HP-positive gastric communities are not uniform, but may contain more than one ecological configuration. In contrast, Gastrotype 3 showed enrichment in *Bacteroides*, *Faecalibacterium*, and *Prevotella*, consistent with a more anaerobic, commensal-like configuration, whereas Gastrotype 4 was characterized by *Streptococcus*, *Veillonella*, *Neisseria*, and *Lactobacillus*, resembling an oral-associated mucosal community. This interpretation is consistent with previous studies showing that *H. pylori* can strongly restructure gastric microbial composition, whereas non-*Helicobacter* gastric dysbiosis is often accompanied by enrichment of oral-associated taxa, particularly in chronic gastric disease and gastric carcinogenesis (Klymiuk et al., 2017; de Leeuw & Duval, 2020; Liu et al., 2022).

### 3.2 Association between gastric types and HP infection status

To evaluate the relationship between gastric type membership and *Helicobacter pylori* (HP) infection status, each gastric type was compared against all remaining samples using Fisher’s exact test. As shown in Table 1, odds ratios (ORs) greater than 1 indicate that the corresponding gastric type is relatively enriched in HP-positive samples, whereas ORs less than 1 indicate relative enrichment in HP-negative samples. In addition, the farther the OR deviates from 1, the stronger the directional association.

**Table 1.** Fisher’s exact test results for the of the four gastric types with HP infection status.

| Gastric Type | HP + (n) | HP - (n) | Total (N) | HP + Proportion | Odds Ratio | P-value | Direction |
| --- | --- | --- | --- | --- | --- | --- | --- |
| 1 | 112 | 28 | 140 | 0.800 | 3.745 | <0.001 | HP+ enriched |
| 2 | 142 | 78 | 220 | 0.645 | 1.495 | 0.028 | HP+ enriched |
| 3 | 22 | 69 | 91 | 0.242 | 0.170 | <0.001 | HP- enriched |
| 4 | 56 | 59 | 115 | 0.487 | 0.602 | 0.019 | HP- enriched |
| Total | 332 | 234 | 566 | 0.587 | NA | NA | NA |

Gastric type 1 was significantly enriched in HP-positive samples, with 112 HP-positive and 28 HP-negative individuals within this cluster (OR = 3.75, *P* < 0.001). Gastric type 2 was also significantly enriched in HP-positive samples (142 HP-positive versus 78 HP-negative; OR = 1.49, *P* = 0.028). In contrast, Gastric type 3 was significantly enriched in HP-negative samples, containing 22 HP-positive and 69 HP-negative individuals (OR = 0.17, *P* < 0.001). Gastric type 4 was likewise significantly associated with HP-negative status (56 HP-positive versus 59 HP-negative; OR = 0.60, *P* = 0.019). Together, these results indicate that the gastric type structure was significantly associated with HP infection status, with Gastric types 1 and 2 showing HP-positive enrichment and Gastric types 3 and 4 showing HP-negative enrichment.

The fact that two distinct gastotypes (Type 1 and Type 2) were both enriched in HP-positive samples suggests that HP-positive gastric communities are not uniform. Rather, HP infection can be associated with at least two different ecological configurations, potentially reflecting differences in host genetics, duration of infection, or co-colonizing microbiota.

Gastotype 3 (Bacteroides-type) resembled a gut-associated anaerobic community, while Gastotype 4 (Streptococcus-type) resembled an oral-associated community. This suggests that HP-negative stomachs can exist in at least two distinct ecological states, consistent with previous observations that non-HP gastric dysbiosis is often accompanied by enrichment of oral taxa (Liu et al., 2022).

### 3.3 Predictive diagnosis of *Helicobacter pylori* infection using gastric type

To establish the baseline predictive performance for *Helicobacter pylori* (HP) infection, six machine-learning classifiers were evaluated, including Logistic Regression, Random Forest, KNN, Gradient Boosting, Linear SVM, and Neural Network. As summarized in Table 2, model performance varied substantially across algorithms. Among the six models, Gradient Boosting achieved the best overall performance, with a mean accuracy of 0.975 (95% CI: 0.973–0.977) and a mean AUC of 0.993 (95% CI: 0.992–0.995). Random Forest showed comparable performance, with a mean accuracy of 0.971 (95% CI: 0.968–0.973) and a mean AUC of 0.990 (95% CI: 0.988–0.992). In contrast, Neural Network exhibited the lowest predictive performance, with a mean accuracy of 0.608 (95% CI: 0.595–0.621) and a mean AUC of 0.646 (95% CI: 0.634–0.658). Logistic Regression and Linear SVM showed intermediate discrimination ability, with mean AUC values of 0.833 and 0.863, respectively, whereas KNN yielded relatively lower performance (mean AUC = 0.795).

**Table 2.** Summary predictive performance of six baseline machine-learning models for HP infection.

| **Model** | **Mean Accuracy** | **95%CI of Accuracy** | **Mean AUC** | **95%CI of AUC** |
| --- | --- | --- | --- | --- |
| Logistic Regression | 0.785 | [0.778, 0.793] | 0.833 | [0.825, 0.842] |
| Random Forest | 0.971 | [0.968, 0.973] | 0.990 | [0.988, 0.992] |
| KNN | 0.712 | [0.705, 0.718] | 0.795 | [0.787, 0.804] |
| Gradient Boosting | 0.975 | [0.973, 0.977] | 0.993 | [0.992, 0.995] |
| Linear SVM | 0.811 | [0.804, 0.818] | 0.863 | [0.855, 0.870] |
| Neural Network | 0.608 | [0.595, 0.621] | 0.646 | [0.634, 0.658] |

The ROC curves further illustrate the differences in model discrimination ability (Table 2 and Figure 4). Consistent with the summary statistics, Random Forest and Gradient Boosting demonstrated the strongest classification performance, with AUC values of 0.990 and 0.989, respectively, whereas Neural Network showed the weakest performance (AUC = 0.638). Logistic Regression, KNN, and Linear SVM produced moderate predictive accuracy, with AUC values of 0.833, 0.795, and 0.862, respectively. Overall, these results indicate that tree-based ensemble methods provided the most effective baseline prediction of HP infection status in the gastric mucosal microbiome dataset.

**Figure 4.**
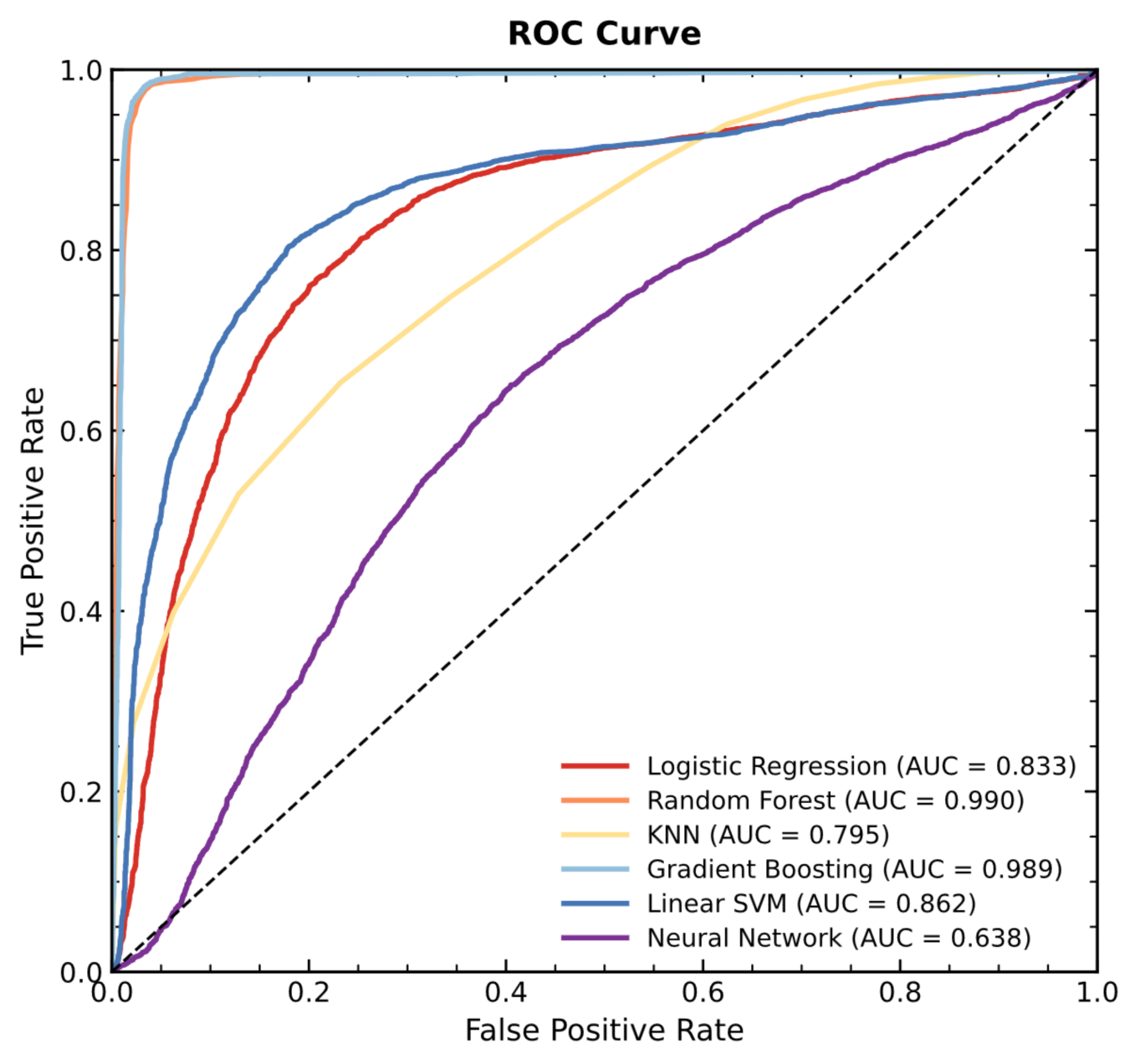
Receiver operating characteristic (ROC) curves of machine learning models for prediction of *Helicobacter pylori* infection status. ROC curves were generated from out-of-fold predictions obtained during repeated stratified cross-validation. Model performances are shown for Logistic Regression, Random Forest, KNN, Gradient Boosting, Linear SVM, and Neural Network classifiers. The diagonal dashed line indicates random classification. The area under the ROC curve (AUC) was highest for Random Forest (0.990) and Gradient Boosting (0.989), followed by Linear SVM (0.862), Logistic Regression (0.833), KNN (0.795), and Neural Network (0.638).

Random Forest and Gradient Boosting clearly outperformed the other classifiers. This result is biologically and methodologically plausible, because microbiome abundance data are high-dimensional, sparse, compositional, and frequently contain nonlinear dependencies among taxa. Tree-based ensemble methods can accommodate such structure more flexibly than linear classifiers and often perform well without requiring extensive feature engineering (Papoutsoglou et al., 2023). In contrast, neural networks generally require larger datasets and more extensive tuning to outperform simpler algorithms on structured microbiome classification tasks (Papoutsoglou et al., 2023). Thus, the ranking observed here is consistent with current experience in microbiome-based machine learning.

### 3.4. Prediction using gastrotype information and VAE-derived representations

To evaluate whether gastric type and VAE-derived features could improve prediction of *Helicobacter pylori* (HP) infection, four Random Forest-based models were compared, including the baseline RF model, Gastric type + RF, VAE + RF, and VAE + Gastric type + RF. As summarized in Table 3, the baseline RF model already achieved excellent predictive performance, with a mean accuracy of 0.972 (95% CI: 0.969–0.974) and a mean AUC of 0.991 (95% CI: 0.989–0.993). Incorporation of gastric type information did not further improve model performance, as the Gastric type + RF model showed nearly identical results, with a mean accuracy of 0.971 (95% CI: 0.969–0.974) and a mean AUC of 0.991 (95% CI: 0.989–0.992).

**Table 3.** Summary predictive performance of four Random Forest-based models for *Helicobacter pylori* infection.

| Model | Mean Accuracy | 95%CI of Accuracy | Mean AUC | 95%CI of AUC |
| --- | --- | --- | --- | --- |
| RF | 0.972 | [0.969, 0.974] | 0.991 | [0.989, 0.993] |
| Gastric type + RF | 0.971 | [0.969, 0.974] | 0.991 | [0.989, 0.992] |
| VAE + RF | 0.653 | [0.644, 0.662] | 0.703 | [0.693, 0.712] |
| VAE + Gastric type + RF | 0.694 | [0.686, 0.702] | 0.765 | [0.757, 0.773] |

The inferior performance of the VAE + RF model may reflect loss of discriminative information during dimensionality reduction. In the prediction branch, the VAE compressed the standardized genus abundance matrix into an eight-dimensional latent representation, and the posterior mean was then used as input for Random Forest classification. Although this representation provides substantial feature compression, it may not preserve all fine-grained signals relevant to HP discrimination that remain available in the original abundance space. In addition, because the VAE was trained with an unsupervised reconstruction objective rather than a classification objective, the learned latent features were not explicitly optimized for prediction.

Within the VAE-based modeling framework, incorporation of gastric type information improved predictive performance. Compared with the VAE + RF model, the VAE + Gastric type + RF model increased mean accuracy from 0.653 to 0.694 and mean AUC from 0.703 to 0.765. This indicates that gastric type provided complementary discriminatory information beyond the VAE-derived latent representation. The ROC curves showed the same pattern (Figure 5). This pattern suggests that gastrotype assignment preserves complementary ecological structure that is not fully retained in a low-dimensional latent embedding. In other words, the VAE representation was useful for revealing community structure.

**Figure 5.**
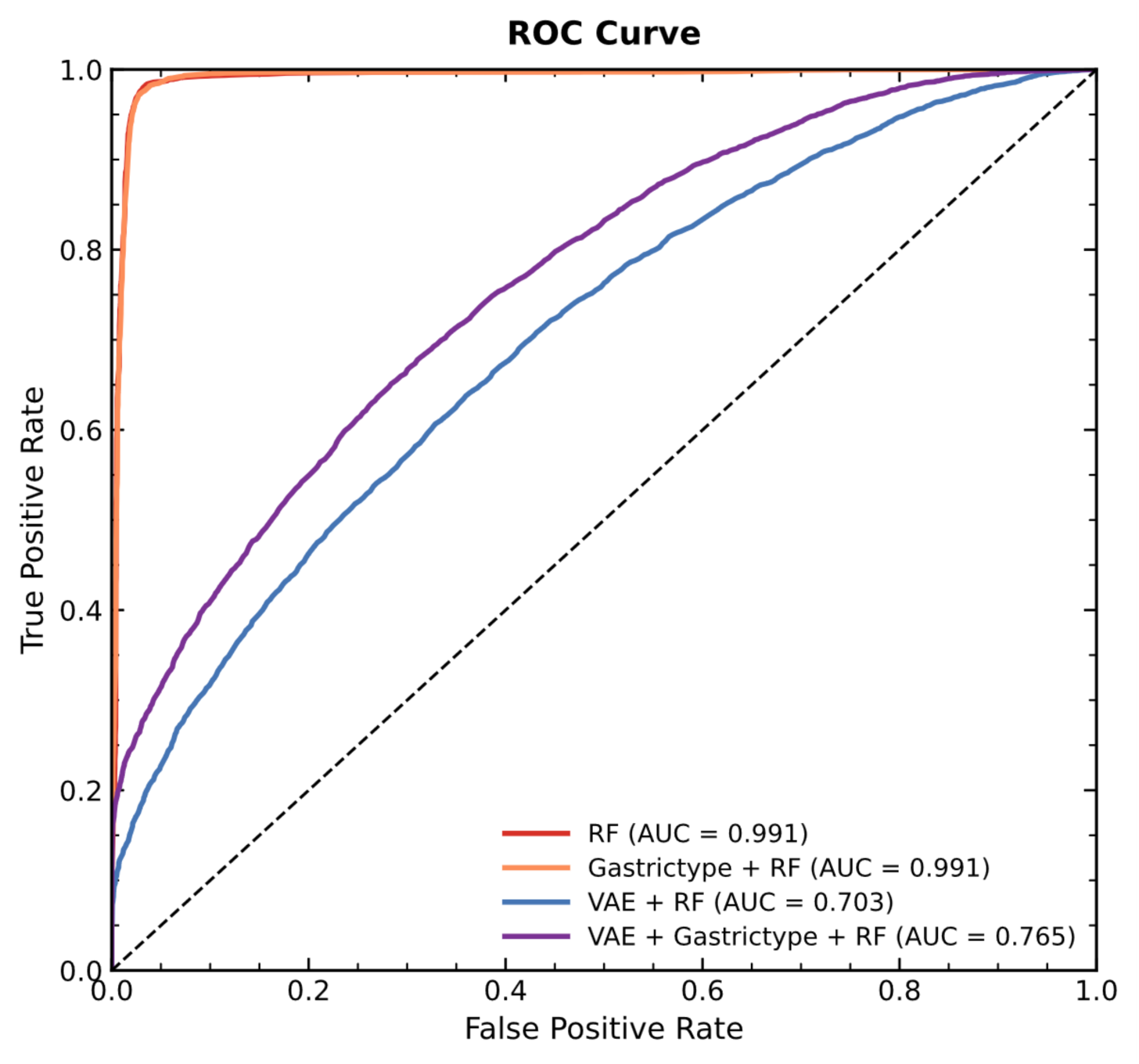
ROC curves comparing the predictive performance of four Random Forest-based models for *Helicobacter pylori* infection.

The improvement of VAE + Gastotype + RF over VAE + RF indicates that gastotype assignment captures complementary ecological structure that is not fully retained in the low-dimensional latent embedding. This suggests that the categorical gastotype labels preserve higher-order community patterns that are partially lost during continuous dimensionality reduction.

## 4. Discussion

In this study, we applied the methodological lessons of the enterotype debate to the gastric mucosal microbiome. Using a variational autoencoder for dimensionality reduction followed by K-means clustering with silhouette-based validation, we identified four recurrent compositional patterns, which we term gastotypes. These gastotypes showed strong and differential associations with *Helicobacter pylori* infection status, and gastotype information provided complementary predictive value beyond raw abundance features. Our findings demonstrate that the rigorous clustering framework developed in response to the enterotype critique can be successfully transferred to a new anatomical site, generating biologically interpretable and clinically relevant community types.

### Comparison to the enterotype literature and methodological contribution

The original enterotype concept proposed that gut microbiomes cluster into three discrete types based on faecal samples (Arumugam et al., 2011). Subsequent critiques demonstrated that enterotype detection depends heavily on methodological choices, that the number of clusters is not fixed, and that absolute validation measures such as the Silhouette index provide weaker support for discrete types than originally claimed (Jeffery et al., 2012; Koren et al., 2013; Knights et al., 2014). A major reconciliation concluded that while the gut microbial composition landscape has local optima or preferred community compositions, discrete boundaries do not exist (Costea et al., 2018).

Our study differs from the original enterotype analysis in two critical ways. First, we determined the optimal number of clusters using the Silhouette index, an absolute validation measure that quantifies cluster cohesion and separation (Rousseeuw, 1987), rather than the Calinski-Harabasz index, a relative measure that assumes clusters exist. Second, we tested a range of K values (2 to 10) and selected K=4 based on the highest mean silhouette score, avoiding the arbitrary selection of three clusters that characterized the original enterotype proposal. The moderate overlap between gastotypes in the VAE latent space (Figure 2) is consistent with the view that these represent recurrent compositional configurations in a continuous space, not discrete, mutually exclusive categories (Costea et al., 2018).

### Biological interpretation of the four gastotypes

The four gastotypes we identified map onto distinct ecological states of the gastric microbiome. Gastotype 1 (Variovorax-type) and Gastotype 2 (Trabulsiella-type) were both significantly enriched in HP-positive samples, but they differed in their taxonomic profiles. Gastotype 1 was dominated by *Helicobacter* (mean relative abundance 0.572) with additional enrichment of *Variovorax* and *Janthinobacterium*. Gastotype 2 showed a more mixed structure with high abundances of *Haemophilus* (0.136), *Helicobacter* (0.106), and *Halomonas* (0.095). This suggests that HP-positive gastric communities are not uniform. Rather, *H. pylori* infection can be associated with at least two distinct ecological configurations, potentially reflecting differences in host genetics, duration of infection, strain variation, or co-colonizing microbiota (Klymiuk et al., 2017; Xiao & Ma, 2022).

Gastotype 3 (Bacteroides-type) and Gastotype 4 (Streptococcus-type) were significantly enriched in HP-negative samples. Gastotype 3 showed high abundances of *Bacteroides* (0.090), *Faecalibacterium* (0.205), and *Prevotella* (0.151), resembling a gut-associated anaerobic community. Gastotype 4 was dominated by *Streptococcus* (0.137), *Lactobacillus* (0.130), and *Prevotella* (0.127), resembling an oral-associated mucosal community. The presence of two distinct HP-negative gastotypes is consistent with previous observations that non-HP gastric dysbiosis is often accompanied by enrichment of oral-associated taxa, particularly in chronic gastric disease and gastric carcinogenesis (Liu et al., 2022; de Leeuw & Duval, 2020). Together, these four gastotypes provide a more nuanced taxonomy of the gastric microbiome than a simple HP-positive versus HP-negative dichotomy.

### Predictive performance and clinical utility

Baseline machine learning models, particularly Random Forest and Gradient Boosting, achieved excellent performance in predicting *H. pylori* infection status, with mean AUC values of 0.990 and 0.993 respectively. This high performance is consistent with prior studies showing that tree-based ensemble methods are well-suited to high-dimensional, sparse, compositional microbiome data, as they naturally handle nonlinear interactions among taxa without requiring extensive preprocessing (Papoutsoglou et al., 2023). The poor performance of the neural network (mean AUC 0.646) is likely attributable to the relatively small sample size (566 samples) relative to the high-dimensional feature space; neural networks typically require larger datasets and extensive hyperparameter tuning to outperform tree-based methods on structured data.

The VAE-based models performed substantially worse than the baseline Random Forest, with VAE + RF achieving a mean AUC of only 0.703. This likely reflects information loss during dimensionality reduction: the VAE compresses the original high-dimensional genus abundance matrix (hundreds of features) into an 8-dimensional latent space, discarding fine-grained discriminatory signals that are preserved in the original feature space and exploited by Random Forest. However, incorporating gastotype information improved the VAE-based model, with VAE + Gastotype + RF achieving a mean AUC of 0.765, compared to 0.703 for VAE + RF alone. This indicates that gastotype assignment captures complementary ecological structure that is not fully retained in the low-dimensional latent embedding. The categorical gastotype labels preserve higher-order community patterns that are partially lost during continuous dimensionality reduction.

### Limitations

Several limitations should be acknowledged. First, our analysis was performed at the genus level due to the limited taxonomic resolution of 16S rRNA sequencing. Species-level or strain-level resolution within key genera such as *Helicobacter*, *Streptococcus*, and *Bacteroides* may reveal additional functional heterogeneity not captured here (Tett et al., 2019). Second, because we pooled multiple public cohorts, the strong predictive performance may partly reflect not only HP-associated biological differences but also stage-specific and cohort-specific structure embedded in the merged dataset. This concern is common in microbiome machine-learning studies and underscores the importance of external validation or stricter leave-one-cohort-out evaluation in future work (Papoutsoglou et al., 2023). Third, our VAE latent space dimensionality (2D for clustering, 8D for prediction) was chosen heuristically rather than optimized systematically; alternative dimensionalities might yield different results. Fourth, the current analysis was performed at the genus level, which improves comparability across datasets but may obscure species-level heterogeneity within key genera. Future studies integrating species-level resolution, cohort-aware validation, and functional profiling may further refine the ecological interpretation of gastric gastotypes. Fifth, our study did not incorporate spatial information about microbial localization within the gastric mucosa. Emerging spatial transcriptomics technologies now enable mapping of host-microbe interactions at sub-micron resolution, and applying these methods to the stomach represents an important future direction (Ntekas et al., 2026; Löstedt et al., 2024).

## Conclusion

In summary, we have identified four recurrent compositional patterns of the gastric microbiome—gastotypes—that are differentially associated with *H. pylori* infection status. By applying the rigorous clustering framework that emerged from the enterotype debate (silhouette-based validation, data-driven determination of K, transparent evaluation of multiple solutions), we provide a robust taxonomy of the gastric microbiome. The four gastotypes map onto distinct ecological states: two HP-enriched configurations (Variovorax-type and Trabulsiella-type) and two HP-negative configurations (Bacteroides-type and Streptococcus-type). While baseline machine learning models achieved excellent predictive performance using raw abundance data, gastotype information provided complementary value when combined with VAE-derived features. Future work integrating species-level resolution, external validation cohorts, and spatial transcriptomic technologies will further refine the clinical utility of gastotypes for predicting gastric disease progression and treatment response.

## Standard Declarations

### Ethical Approval

Ethical approval is not applicable since the study only analyzed the data from public domain.

### Data Accessibility Statement

All data reanalyzed in this study are publicly available, with access details provided in Table S1. The computational codes are also publicly accessible and can be found in the respective method papers cited in the Materials and Methods section.

## References

1. Arumugam, M., Raes, J., Pelletier, E., Le Paslier, D., Yamada, T., Mende, D. R., Fernandes, G. R., Tap, J., Bruls, T., Batto, J. M., Bertalan, M., Borruel, N., Casellas, F., Fernandez, L., Gautier, L., Hansen, T., Hattori, M., Hayashi, T., Kleerebezem, M., Kurokawa, K., Leclerc, M., Levenez, F., Manichanh, C., Nielsen, H. B., Nielsen, T., Pons, N., Poulain, J., Qin, J., Sicheritz-Ponten, T., Tims, S., Torrents, D., Ugarte, E., Zoetendal, E. G., Wang, J., Guarner, F., Pedersen, O., de Vos, W. M., Brunak, S., Dore, J., Weissenbach, J., Ehrlich, S. D., & Bork, P. (2011). Enterotypes of the human gut microbiome. Nature, 473(7346), 174–180.

2. Coker, O. O., Dai, Z., Nie, Y., Zhao, G., Cao, L., Nakatsu, G., Wu, W. K., Wong, S. H., Chen, Z., Sung, J. J. Y., & Yu, J. (2018). Mucosal microbiome dysbiosis in gastric carcinogenesis. Gut, 67(6), 1024–1032.

3. Costea, P. I., Hildebrand, F., Arumugam, M., Backhed, F., Blaser, M. J., Bushman, F. D., de Vos, W. M., Ehrlich, S. D., Fraser, C. M., Hattori, M., Huttenhower, C., Jeffery, I. B., Knights, D., Lewis, J. D., Ley, R. E., Ochman, H., O’Toole, P. W., Quince, C., Relman, D. A., Shanahan, F., Sunagawa, S., Wang, J., Weinstock, G. M., Wu, G. D., Zeller, G., Zhao, L., Raes, J., Knight, R., & Bork, P. (2018). Enterotypes in the landscape of gut microbial community composition. Nature Microbiology, 3(1), 8–16.

4. de Leeuw, M. A., & Duval, M. X. (2020). The presence of periodontal pathogens in gastric cancer. Exploratory Research and Hypothesis in Medicine, 5(3), 87–96.

5. Higgins, I., Matthey, L., Pal, A., Burgess, C., Glorot, X., Botvinick, M., Mohamed, S., & Lerchner, A. (2017). β-VAE: Learning basic visual concepts with a constrained variational framework. In Proceedings of the International Conference on Learning Representations (ICLR). https://openreview.net/forum?id=Sy2fzU9gl

6. Jeffery, I. B., Claesson, M. J., O’Toole, P. W., & Shanahan, F. (2012). Categorization of the gut microbiota: enterotypes or gradients? Nature Reviews Microbiology, 10(9), 591–592.

7. Kingma, D. P., & Welling, M. (2014). Auto-encoding variational Bayes. In Proceedings of the International Conference on Learning Representations (ICLR). https://arxiv.org/abs/1312.6114

8. Klymiuk, I., Bilgilier, C., Stadlmann, A., Thannesberger, J., Kastner, M.-T., Högenauer, C., Püspök, A., Biowski-Frotz, S., Schrutka-Kölbl, C., Thallinger, G. G., & Steininger, C. (2017). The human gastric microbiome is predicated upon infection with Helicobacter pylori. Frontiers in Microbiology, 8, 2508.

9. Knights, D., Ward, T. L., McKinlay, C. E., Miller, H., Gonzalez, A., McDonald, D., & Knight, R. (2014). Rethinking “enterotypes.” Cell Host & Microbe, 16(4), 433–437.

10. Koren, O., Knights, D., Gonzalez, A., Waldron, L., Segata, N., Knight, R., Huttenhower, C., & Ley, R. E. (2013). A guide to enterotypes across the human body: meta-analysis of microbial community structures in human microbiome datasets. PLOS Computational Biology, 9(1), e1002863.

11. Ling, Z., Shao, L., Liu, X., Cheng, Y., Yan, C., Mei, Y., Ji, F., & Liu, X. (2019). Regulatory T Cells and Plasmacytoid Dendritic Cells Within the Tumor Microenvironment in Gastric Cancer Are Correlated With Gastric Microbiota Dysbiosis: A Preliminary Study. Frontiers in Immunology, 10, 533. 10.3389/fimmu.2019.00533

12. Liu, D., Zhang, R., Chen, S., Sun, B., & Zhang, K. (2022). Analysis of gastric microbiome reveals three distinctive microbial communities associated with the occurrence of gastric cancer. BMC Microbiology, 22(1), 184. 10.1186/s12866-022-02594-y

13. Löstedt, B., Stražar, M., Xavier, R., Regev, A., & Vickovic, S. (2024). Spatial host-microbiome sequencing reveals niches in the mouse gut. Nature Biotechnology, 42(9), 1394–1403.

14. Malfertheiner, P., Megraud, F., O’Morain, C. A., Gisbert, J. P., Kuipers, E. J., Axon, A. T., Bazzoli, F., Gasbarrini, A., Atherton, J., Graham, D. Y., Hunt, R., Moayyedi, P., Rokkas, T., Rugge, M., Selgrad, M., Suerbaum, S., Sugano, K., & El-Omar, E. M. (2017). Management of Helicobacter pylori infection—the Maastricht V/Florence consensus report. Gut, 66(1), 6–30.

15. Ma Z.S. & Li LW. (2017) Quantifying the human vaginal community state types (CSTs) with the species specificity index. Peer J, 5: e3366.

16. Ma Z.S. (2023). A new hypothesis on BV etiology: dichotomous and crisscrossing categorization of complex versus simple on healthy versus BV vaginal microbiomes. mSystems, 8(5), e0004923. 10.1128/msystems.00049-23

17. McCallum, G., & Tropini, C. (2024). The gut microbiota and its biogeography. Nature Reviews Microbiology, 22(2), 105–118.

18. Ntekas, I., Takayasu, L., McKellar, D. W., Grodner, B. M., Holdener, C., Schweitzer, P., Park, Y. S., Sauthoff, M., Shi, Q., Brito, I. L., & De Vlaminck, I. (2026). Spatial transcriptomics maps host–gut microbiome biogeography at high resolution. Nature Microbiology. Advance online publication. 10.1038/s41564-026-02286-7

19. Papoutsoglou, G., Tarazona, S., Lopes, M. B., Klammsteiner, T., Ibrahimi, E., Eckenberger, J., Novielli, P., Tonda, A., Simeon, A., Shigdel, R., Béreux, S., Vitali, G., Tangaro, S., Lahti, L., Temko, A., Claesson, M. J., & Berland, M. (2023). Machine learning approaches in microbiome research: challenges and best practices. Frontiers in Microbiology, 14, 1261889. 10.3389/fmicb.2023.1261889

20. Park, C. H., Lee, A. R., Lee, Y. R., Eun, C. S., Lee, S. K., & Han, D. S. (2019). Evaluation of gastric microbiome and metagenomic function in patients with intestinal metaplasia using 16S rRNA gene sequencing. Helicobacter, 24(1), e12547. 10.1111/hel.12547

21. Qiao, Y.T., & Ma, Z. S. (2026). A VAE-based methodology for deep enterotyping and Parkinson’s disease diagnosis. medRxiv preprint doi: 10.64898/2026.03.17.26348604

22. Rousseeuw, P. J. (1987). Silhouettes: A graphical aid to the interpretation and validation of cluster analysis. Journal of Computational and Applied Mathematics, 20, 53–65. DOI: 10.1016/0377-0427(87)90125-7

23. Siezen, R. J., & Kleerebezem, M. (2011). The human gut microbiome: are we our enterotypes? Microbial Biotechnology, 4(5), 550–553.

24. Tett, A., Huang, K. D., Asnicar, F., Fehlner-Peach, H., Pasolli, E., Karcher, N., Armanini, F., Manghi, P., Bonham, K., Zolfo, M., De Filippis, F., Magnabosco, C., Bonneau, R., Lusingu, J., Amuasi, J., Reinhard, K., Rattei, T., Boulund, F., Engstrand, L., Zink, A., … Segata, N. (2019). The Prevotella copri Complex Comprises Four Distinct Clades Underrepresented in Westernized Populations. Cell Host & Microbe, 26(5), 666–679.e7. 10.1016/j.chom.2019.08.018

25. Wang, Z., Gao, X., Zeng, R., Wu, Q., Sun, H., Wu, W., Zhang, X., Sun, G., Yan, B., Wu, L., Ren, R., Guo, M., Peng, L., & Yang, Y. (2020). Changes of the Gastric Mucosal Microbiome Associated With Histological Stages of Gastric Carcinogenesis. Frontiers in Microbiology, 11, 997. 10.3389/fmicb.2020.00997

26. Xiao, W. M., & Ma, Z. S. (2022). Influences of Helicobacter pylori infection on diversity, heterogeneity, and composition of human gastric microbiomes across stages of gastric cancer development. Helicobacter, e12899. 10.1111/hel.12899

